# State-specific Projection of COVID-19 Infection in the United States and Evaluation of Three Major Control Measures

**DOI:** 10.1101/2020.04.03.20052720

**Authors:** Shi Chen, Qin Li, Song Gao, Yuhao Kang, Xun Shi

## Abstract

Most models of the COVID-19 pandemic in the United States do not consider geographic variation and spatial interaction. In this research, we developed a travel-network-based susceptible-exposed-infectious-removed (SEIR) mathematical compartmental model system that characterizes infections by state and incorporates inflows and outflows of interstate travelers. Modeling reveals that curbing interstate travel when the disease is already widespread will make little difference. Meanwhile, increased testing capacity (facilitating early identification of infected people and quick isolation) and strict social-distancing and self-quarantine rules are most effective in abating the outbreak. The modeling has also produced state-specific information. For example, for New York and Michigan, isolation of persons exposed to the virus needs to be imposed within 2 days to prevent a broad outbreak, whereas for other states this period can be 3.6 days. This model could be used to determine resources needed before safely lifting state policies on social distancing.

## Introduction

The Coronavirus disease (COVID-19) is an ongoing pandemic that poses a global threat. As of March 26, 2020, more than 520,000 cases of COVID-19 have been reported in over 200 countries and territories, resulting in approximately 23,500 deaths^1–9^. In the United States, the first known positive case was identified in Washington state on January 20, 2020^10^. By March 26, the epidemic had been rapidly spreading across many communities and present in all 50 states, plus the District of Columbia; the total number of confirmed cases in the United States rose to 78,786 with 1,137 deaths.

To combat the spread of COVID-19, the government has taken actions in various dimensions, including banning or discouraging domestic and international travels, announcing stay-at-home orders to curb non-essential interactions for reducing transmission rate, and urging commercial laboratories to increase test capacity. To curb traveling, on January 31, the United States government announced travel restrictions on travelers from China; on February 29, it announced travel ban against Iran and advised travel with caution to Europe^11^; on March 11, it announced travel restrictions on most of European countries. To reduce human-interactions, on March 13, a national emergency was declared; as of March 28, 39 states had issued either statewide or regionally stay-at-home or shelter-in-place order, requiring residents to stay indoors except for essential activities. To increase test capacities, on February 4, the United States Food and Drug Administration (FDA) approved the United States Centers for Disease Control and Prevention (CDC)’s test, which was later to be proved inconclusive^12^; on February 29, the FDA relaxed its rules for some laboratories, allowing them to start testing before the agency granting its approvals; on March 27, FDA issued an Emergency Use Authorization to a medical device maker, the Abbott Labs, for the use of a coronavirus test that delivers quick testing results^13^.

So far, since there is no treatment or vaccine for SARS-COV-2 available, these actions have been taken largely based on classic non-pharmaceutical epidemic controls. Works on evaluating similar measures in other countries, especially China, started to emerge^7,14,15^. For example, the effect of travel restriction on delaying the virus spread in China has been reported^5,16^. However, it is still unclear what control and intervention measures would have actual effect, especially to what extent, on abating the spread of COVID-19 in the United States. As the United States has very different political, administrative, social, pubic health and medical systems, as well as culture from China, this remains to be a critical question to address, especially considering that some measures and policies come with extremely high economic and societal costs.

There have been numerous modeling works projecting or predicting the trend of the COVID-19 pandemic regionally or globally^17,18^. Most of the works apply a global model to the entire study area, either a region, a country, or the entire globe. Rarely the variation of different parts within one area and the interactions among those parts are taken into consideration. However, a country like the United States features diversity in all aspects. On the one hand, the overall situation of the entire country is a result emerging from local situations and their interactions, and thus, ignoring the local interactions can hardly lead to a high-quality overall model; on the other hand, as all interventions and policies finally have to be adapted to the local situation, a localized modeling will be much more relevant to the real-world practices. Spatially and network-related epidemic models can describe the geographical spread of viral dynamics^7,19–21^. Recent studies have shown the importance of incorporating timely human mobility patterns derived from mobile phone big data and global flight networks into the epidemiology modeling process and in public health studies^5,7,22–30^. Without accurate models that incorporate human mobility patterns and spatial interactions^26,27^, it is rather challenging to quantify the sensitivity of parameters, and using the linkage to real practices to make sensible policy suggestions.

Accordingly, the core of the study is twofold. First, to localize the modeling, we developed a travel-network-based susceptible-exposed-infectious-removed (SEIR) mathematical compartmental model system that simultaneously characterizes the spatiotemporal dynamics of infections in 51 areas (50 states and the District of Columbia). Each state or district has its own model, and all models simultaneously take into account inflows and outflows of interstate travelers.

Second, to improve the practical relevance, we chose to use three parameters that can directly correspond to possible practical means to discover, combat, and control the spread of the disease, and quantify their impact on the final output of the model. The three parameters include: 1) the transmission rate *b*, which corresponds to the local social-distancing enforcement, e.g., the stay-home order; 2) the detection and reporting rate *r*, which corresponds to the testing capacity; and 3) the travel ratio *α*_*t*_, which corresponds to the ratio of interstate travel volume compared to that of 2019 during the same period.

The modeling is a dynamic projection process (see the ‘methods’ section). We employed daily and state-specific historical data to incrementally calibrate the model, and then used the calibrated model to predict future scenarios under different non-pharmaceutical control and intervention measures. During this process, we ran data assimilation methods to identify parameter values that optimally fit the current situation (see more details in the methods and supplementary material). To project into the future, we set different values for the parameters to create different control and intervention scenarios, and then ran the simulation to see their impact on the model results. The final output of the model is the total number of confirmed cases in a state on a particular day. The current strategy in the United States is to isolate people who have the symptoms of COVID-19. An ideal scenario is to have an 100% reporting rate, i.e., every infected case gets confirmed and thus isolated quickly. Another ideal setting is to have everyone who was in contact with the infected gets identified and isolated quickly as well. Our model incorporated these considerations and examined such direct isolation of the exposed compartment in detail. We particularly investigated the impact of quickness of such actions through mathematical modeling and scenario analysis.

A notable result from our modeling is that the impact of interstate travel restriction on the model output is modest. This can be explained by that when the disease has already widespread in all states, the relatively small number of cases in the travelers will cause little difference to the local situation, compared with the effects of local social-distancing and isolation rules and the increase of testing capacity.

## Results

Fig. 1 shows the effect on spatiotemporal dynamics of infectious population across states by setting the coefficients at different configurations. An interactive map-based scenario simulation web dashboard is also available at https://geods.geography.wisc.edu/covid19/us_model. We set *r* = 1 − *α*_*r*_(1 − *r*_0_) and *b* = *α*_*b*_*b*_0_, where *r*_0_ and *b*_0_ are the report and transmission rate as of March 20, 2020 using data assimilation fitting result. By decreasing *α*_*r*_ from 1 to 0, we increase the report rate from the original *r*_0_ to 1, and by decreasing *α*_*b*_ we decrease the transmission rate. Most states, except a few such as NY, MI, and CA, see drastic improvement when the transmission rate is decreased and the testing(reporting) rate is increased, but the reduction of interstate traffic alone is not as effective. Our modelling reveals that once the epidemic in an area has reached a certain stage, the difference that can be caused to the local situation by the relatively small number of imported cases due to the interstate travel is insignificant. According to our modeling, all states in the United States have reached that stage. Therefore, as long as those travelers follow the social-distancing rules and the local government provides sufficient testing capacity, there is no apparent urge to curb interstate travel. This is in line with the finding in^16,28^, in which the authors projected the pick up of the spreading in other parts of China outside of Wuhan with about 3 days delay, and in the world outside China within a 2-3 weeks of delay, assuming no further screening is in place. Different from China where the city of Wuhan is clearly the epicenter of the COVID-19 outbreak and the travel ban quickly gets the rest of China under control, most of the states in the United States have already had signs of community spread by March 20, 2020^31^, and banning other states will hardly make much difference to the local situation. In addition, Fig. 2 shows the corresponding prediction time series of infectious population in top 15 states under two scenarios (see also Fig. S14): (A) the reported rate and the transmission rate remained unchanged as of March 20, 2020, with *α*_*r*_ = *α*_*b*_ = 1, in which most states will continue their exponential growth before reaching their peak; (B) with *α*_*r*_ = *α*_*b*_ = 0.1, that is, when the transmission rate *b* is much smaller and the reported rate *r* is much higher (closer to 1), we can “flatten the curve” on the virus (i.e., reducing the spread of the virus).

**Figure 1.**
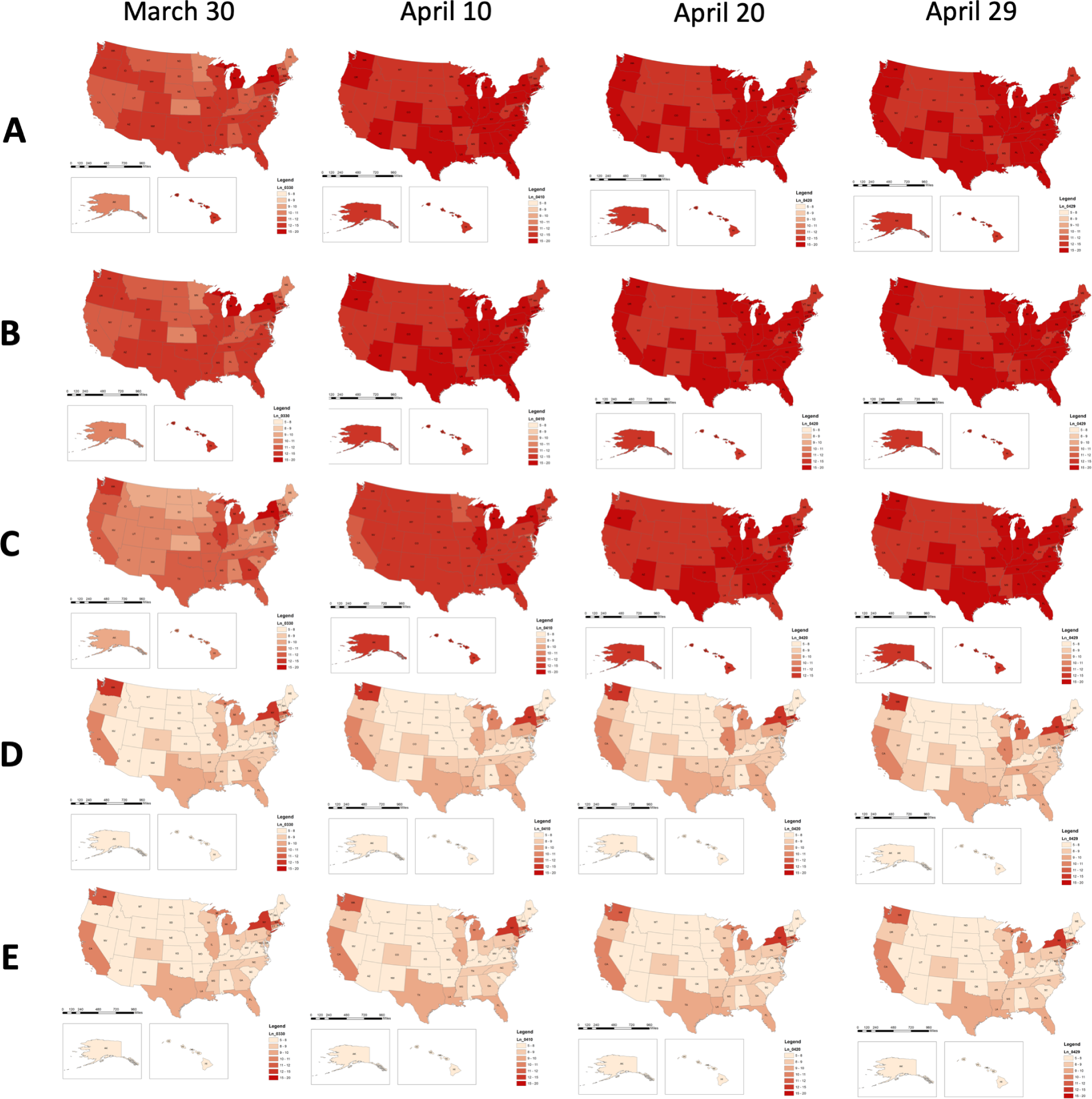
The spatiotemporal distribution of predicted infected population (in natural logarithm scale) across all states under different simulation scenarios: (A) *α*_*r*_ = 1 and *α*_*b*_ = 1, i.e., all parameters took the values of the initial configuration, obtained through data assimilation method using the numbers of confirmed cases during March 1 – March 20, 2020; (B) the travel flow was reduced to *α*_*t*_ = 0.05, while other parameters values remained unchanged; (C) *α*_*r*_ = 0.1 and *α*_*b*_ = 1; (D) *α*_*r*_ = 1 and *α*_*b*_ = 0.1; (E) *α*_*r*_ = 0.1, *α*_*b*_ = 0.1. In the simulations, the transmission rate was set to be *b* = *α*_*b*_*b*_0_ and the reporting rate was set to be *r* = 1 − *α*_*r*_(1 − *r*_0_). Where *r*_0_ and *b*_0_ were the reporting rate and the transmission rate on March 20, 2020, which are inferred from the data assimilation step.

**Figure 2.**
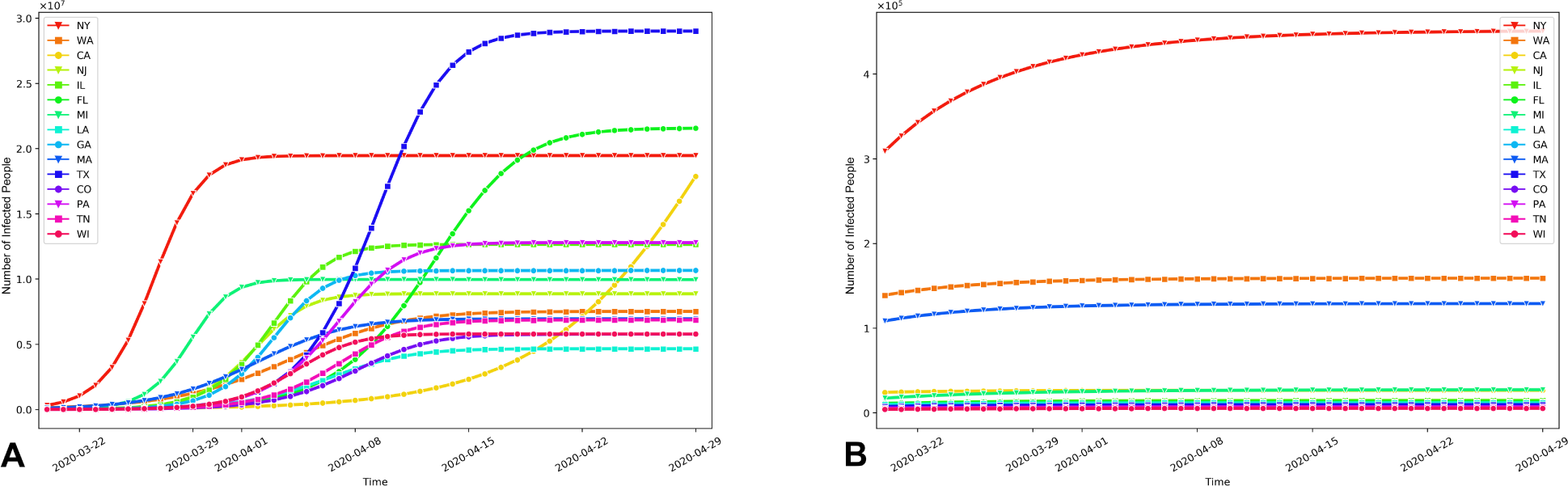
The prediction time series of the total infected population in the 15 most affected states under two scenarios: (A) *α*_*r*_ = *α*_*b*_ = 1, i.e., both the reported rate and the transmission rate remained unchanged; (B) *α*_*r*_ = *α*_*b*_ = 0.1, i.e., the transmission rate *b* was smaller and the reported rate *r* was larger (closer to 1) as *r* = 1 −*α*_*r*_(1 −*r*_0_).

We further investigate the effect of increased testing capacity and report rate. As shown in Figure 3a, most states see drastic improvement when the report rate increases. All states, by April 29, see monotonically exponential reduction of infections. The impact is strong in states such as MA, AZ, FL, and OR, but relatively weak in states such as NY, MI and IL. In Figure 3b, we study the effect of *α*_*r*_ and *α*_*b*_ on the basic reproduction rate *R*_*e*_ in NY (see other states in Fig. S15). It can be seen that merely raising the report rate cannot fully make *R*_*e*_ *<* 1. To mitigate the spread of COVID-19 in these states, a proactive approach needs to be taken, and quick detection and isolation of the exposed population need to be in place instead of being delayed until the onset of the symptoms. This measure can prevent the exposed population from potentially infecting other susceptible people. In Figure 3c, we plot the increase of infections in terms of *D*_*q*_ (i.e., the temporal lag in putting a person into quarantine) for the states that are sensitive to change of *D*_*q*_, including NY, NJ, IL, GA, MI, CO, WI, LA, TX, PA, MA, and TN. The longer one waits to inform and isolate the exposed population, the more infected people one observes. For example, there is a sharp transition for NY and MI. If the average detection and isolation time is more than 2 days, the total number of infections will significantly increase.

**Figure 3.**
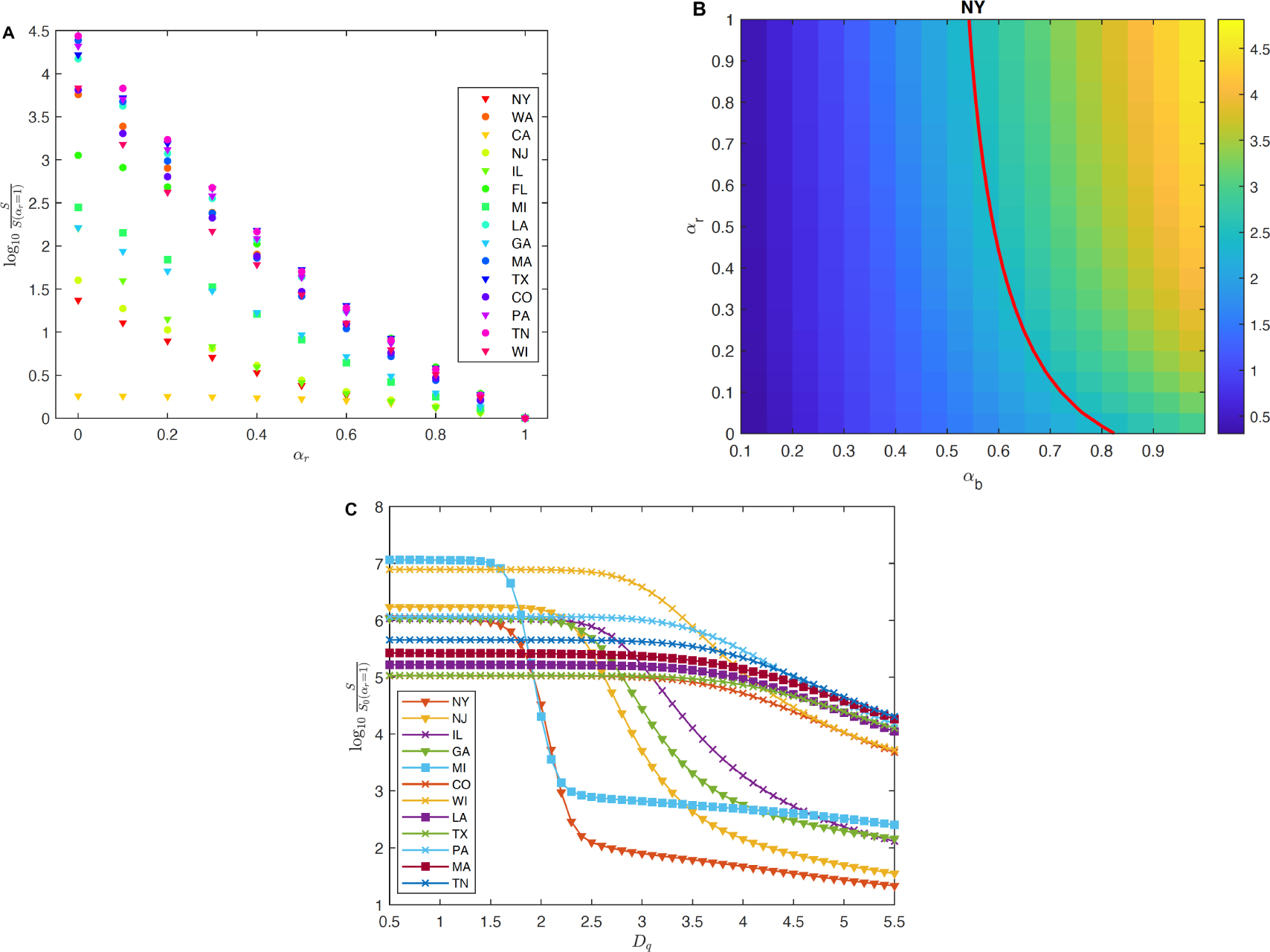
(A) Susceptible population (*S*) on April 29, 2020 as a function of *α*_*r*_. *S*(*α*_*r*_ = 1) is the susceptible population on April 29 computed with the report rate set as the original report rate inferred from the data assimilation step. In all states, *S* increases as *α*_*r*_ decreases, meaning that more people stay unaffected when a higher report is enacted. (B) *R*_*e*_, the basic reproduction number, on April 29 for different *α*_*b*_ and *α*_*r*_ in NY. The red line is the level set *R*_*e*_ = 1. It can be seen that increasing the reported rate helps diminish the reproductive number, but cannot reduce *R*_*e*_ under 1 if the original transmission rate *b*_0_ is applied; (C) Susceptible population on April 29 for different *D*_*q*_. *S*(*α*_*r*_ = 1) is the same as in (A). *S* significantly depends on the period from expose to quarantine.

The results again showed the importance of sufficient testing and strong transmission-intervention measures such as social distancing and self-quarantine policy^32^. These policies can help quickly identify the source of infection and isolate them before they infect the remaining population. This measure presumably comes with a lower economical cost.

We finally investigate the stability of our statements on the parameters chosen in the model. There are a number of parameters in the model that are determined according to medical studies and thus necessarily contain ambiguity. One parameter, *γ*, is especially hard to be set at a particular value due to the lack of medical evidence. This parameter reflects the level of infectiousness of the “exposed” compartment, a population that is presymptomatic. Recent studies indicate that presymptomatic patients seem to be more infectious than patients who have symptoms on site^33^. We therefore run our model with different values of *γ* to identify the significance of this particular parameter. Our numerical result suggests that within a moderate range of *γ*, our conclusions still stand true. In particular, as shown in Figure 4, by setting the “exposed” compartment being more infectious than the “infected” compartment, the numerical solution shows the same trend. We still observe that, with a higher report rate, the number of non-infected population exponentially increases (i.e., less people would get infected), and when a proactive approach is taken, meaning that the “exposed” compartment gets quickly separated from the rest of the population, the non-infected population drastically increases as *D*_*q*_, the delay of the separation time, gets shortened. This means that the dependence of our conclusion on the parameter *γ* is stable, and the above statements are consistent.

**Figure 4.**
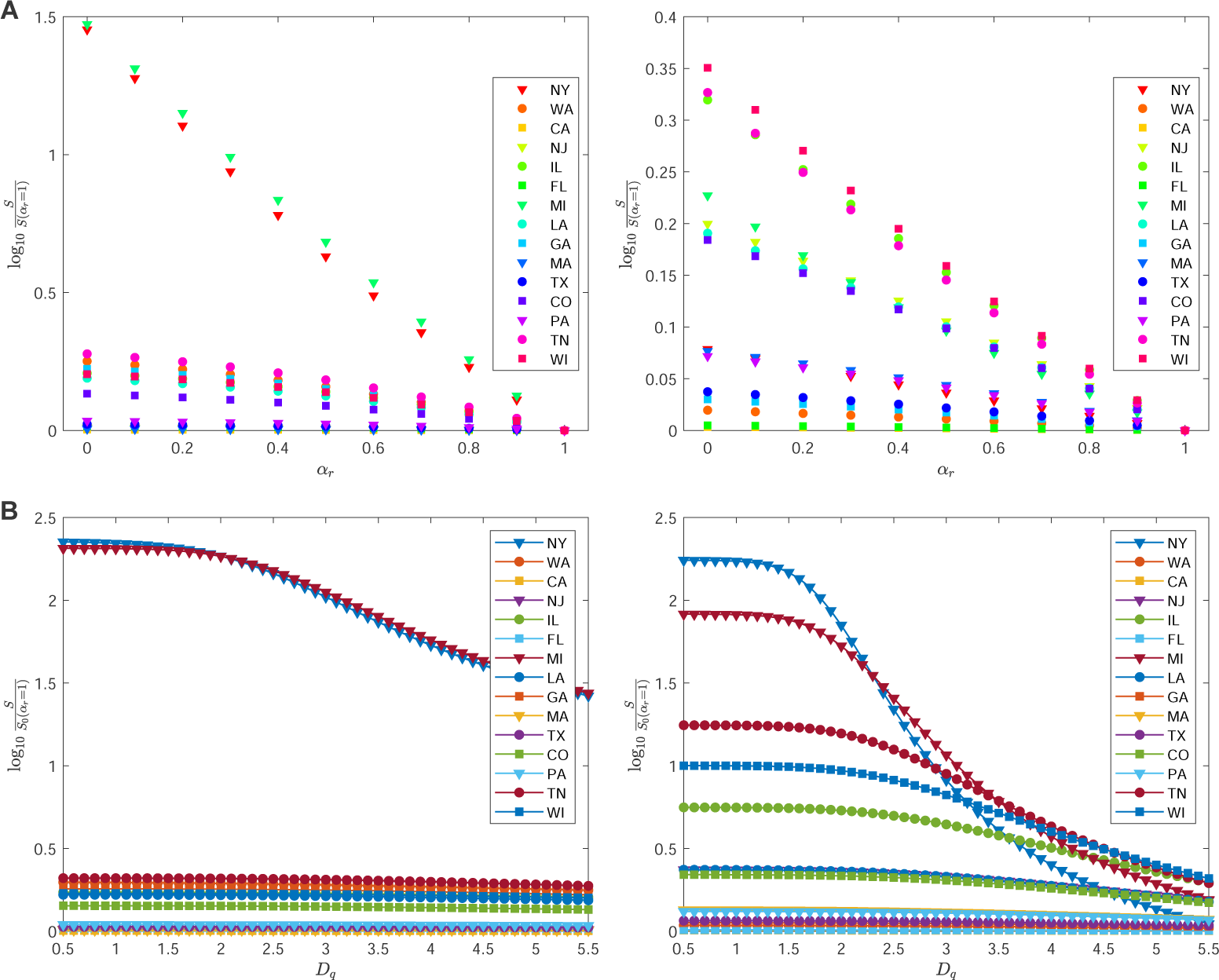
(A) Susceptible population (*S*) on April 4, 2020 as a function of *α*_*r*_. The panels on the left and on the right are results from *γ* = 0.5 and *γ* = 1.5, respectively. For both *γ, S* increases as *α*_*r*_ decreases, meaning that more people stay unaffected when a higher report is enacted. (B) Susceptible population on April 4, 2020 for different *D*_*q*_ and different values of *γ*. For those states whose susceptible population is much smaller than their total population due to a high infection rate (such as in NY), *S* significantly depends on *D*_*q*_ for both *γ <* 1 and *γ >* 1.

We should emphasize that in our simulation, we do not differentiate patients with severe or mild symptoms. A more dedicated numerical experiment that separates the two categories could potentially give more detailed information. For example, in another agent-based modeling study^34^, researchers consider patients with mild to severe symptoms to evaluate the impacts of the timing of social distancing and adherence level on COVID-19 confirmed cases.

## Discussion and conclusion

Modeling and analyzing the spread of COVID-19, and assessing the effect of various policies could be instrumental to national and international agencies for health response planning^5,8,15–17,32^. We show that the effect of interstate travel reduction is at most modest in the United States when the outbreak has already widespread in all states. On the other hand, we need to impose strong transmission-reduction intervention and increased testing capacity and report rate to contain the spread of virus. The result is based on mathematical and statistical analyses of transmission control measures and in agreement with previous findings^2,3,5,14–16^, suggesting that the effect of travel ban at a later stage of the outbreak is rather modest. This is also in line with the fact that the outbreaks still occurred in Europe even upon the strong travel ban on the earlier epicenter of Wuhan and its surrounding cities in China. We also quantitatively show that the transmission-reduction intervention such as policies on the social-distancing and shelter-in-place rules, and the increase of testing rate, which facilitates immediate isolation upon exposure, will significantly reduce the total infected population. Such effect is mostly visible for the states of NY, NJ, MI, and IL. Particularly, our modeling results show that for states such as NY and MI, to achieve an optimal infection reduction, a more proactive approach needs to be taken to quickly identify the exposed population and isolate them within two days of exposure in order to ensure the infection reduction. The result is in agreement with previous findings^7,8^.

We do need to emphasize that the model itself does not distinguish different ways of traveling across states. Indeed, if the interstate travel is conducted mostly through transiting through busy airports and train stations, and the social-distancing policy is not strictly imposed, then the high population density at these places will bring up the transmission rate *b* locally in space and time, leading to a higher infection rate. This is a severe consequence, but it should not be counted as the direct result of relaxing travel restrictions.

Moving forward, we estimate that the decline in travel has a modest effect on the mitigation of the pandemic. We need a stronger transmission-reduction intervention and increased detection and report rate in place to prevent the further spread of the virus. The results could potentially be used to design an optimal containment scheme for mitigating and controlling the spread of COVID-19 in the United States.

## Methods

The mathematical model that simulates the spatiotemporal dynamics of state-level infections in the United States is a modified travel-network-based SEIR compartmental model in epidemiology by taking into account the variation of the 51 administrative units and their interactions^14,35–37^. It consists of 51 ordinary differential equation (ODE) systems, with each one characterizing the evolution of susceptible (*S*), exposed (*E*), reported (*I*), unreported (*U*) and removed (*R*) cases per state (Fig. S1 and see more details in the supplementary material). The 51 ODE systems are then coupled through the state-to-state travel network flows (see Fig. S2) that were extracted from the aggregated SafeGraph mobility data and weighted by *α*_*t*_^38, 39^. Unlike most other models, we also incorporate the potential asymptomatic transmission. This makes the derivation of the basic reproduction number *R*_0_ different. Besides, each ODE system also includes two unknown parameters: the transmission rate (*b*) and the report rate for each state (*r*). The unknown parameters are inferred based on the total number of confirmed cases in each state for the period of March 1-March 20, 2020. The source of infection case data is the Center For Systems Science and Engineering at the Johns Hopkins University^9^.

The parameters and model specification are defined as follows:

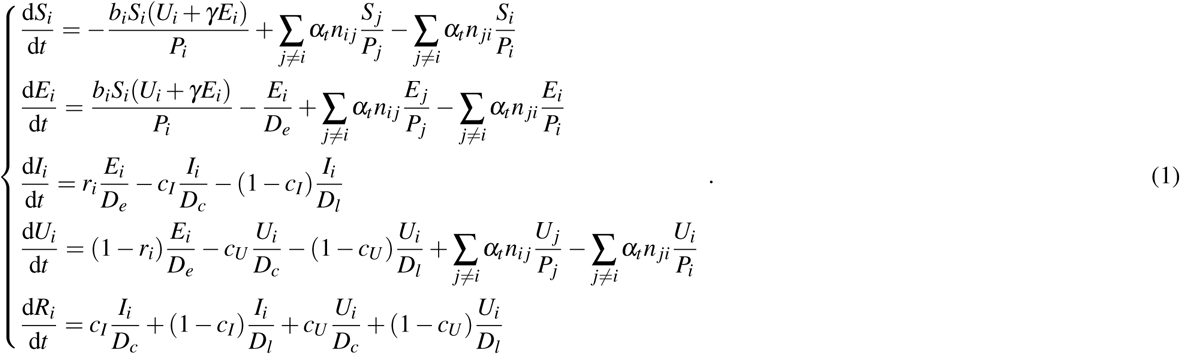

The ODE system is equipped with the following initial data (*t* = 0 standing for March 1, 2020):

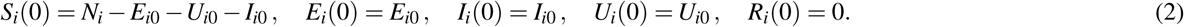

In the equation, the unit for *t* is one day. *N*_*i*_(*t*) is the total population of state *i* at time *t*, and *P*_*i*_ = *S*_*i*_ + *E*_*i*_ +*U*_*i*_ is the free population. *n*_*i j*_ is the number of inflow from state *j* to state *i. b*_*i*_ and *r*_*i*_ are the transmission rate and reporting rate of state *i. c*_*I*_ (*c*_*U*_, resp.) is the proportion of positive cases that show critical condition for *I* (unreported cases *U*, resp.). *D*_*e*_ is the latent period. *D*_*c*_ and *D*_*l*_ are the infectious periods of critical cases and mild cases. *α*_*t*_ is a parameter to tune the traffic flow.

We emphasize two main differences in modeling compared with existing literature. In^7^, the authors study the inter-city traffic and its impact on the spreading of COVID-19 in China. The situation in China and that in the US are very different. In China, the epicenter is clear: the city of Wuhan, Hubei province, and the outbreak starts mid-January, 2020. The COVID-19 outbreak in the US, however, is multi-sourced. The consequence is that in the model in^7^, the initial condition for cities excepts Wuhan is clear: the latent, the reported and the unreported cases are all zero. In this model, however, the initial conditions *E*_*i*0_ are unclear for all states; Another big difference is, according to clinical findings, the latent cases also have the potential of transmitting the virus, and thus we add the interaction of *E*_*i*_ with *S*_*i*_ into the increment of *E*_*i*_^7,40,41^.

The unknown parameters and state variables in the equation set are

* *b*_*i*_: the transmission rate with non-informative prior range [1, 1.5];
* *r*_*i*_: the report rate with non-informative prior range [0.1, 0.3];
* *E*_*i*0_: the data for the latent population with non-informative prior range [0, 500].
* *U*_*i*0_: the initial data for the unreported population with non-informative prior range [0, 200].
* *S*_*i*0_: the initial data for the susceptible population defined by *N*_*i*_ −*E*_*i*0_ −*I*_*i*0_ −*A*_*i*0_.

Other parameters are:

*γ*: the transmission ratio between unreported and latent. In the simulation we set it to be 0.5;

*D*_*c*_: the average duration of infection for critical cases. We assume *D*_*c*_ = 2.3 days^42^.

*D*_*e*_: the average latent period. According to^43^, *D*_*e*_ = 5.2 days.

*D*_*l*_: the average duration of infection for mild cases. We assume *D*_*l*_ = 6 days.

*α*_*t*_: the ratio of interstate travel volume compared to that of 2019 during the same period. The travel flow information *n*_*i j*_ was extracted from the SafeGraph mobility data, and we set *α*_*t*_ = 0.5 to represent the travel reduction situation observed in the year of 2020.

*c*_*I*_: proportion of critical cases among all reported cases. We choose *c*_*I*_ = 0.1

*c*_*U*_: proportion of critical cases among all unreported cases. We assume *c*_*A*_ = 0.2.

There is an essential assumption made in the model: the homogeneity in the population. It means that the traffic flow is a good representation of the total population without considering their demographic and socioeconomic characteristics. The susceptible, exposed, and unreported move in and out of states at the same rate. This explains the 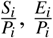 and 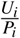 terms in the *S*_*i*_*/E*_*i*_*/U*_*i*_ equation.

The effective reproductive number *R*_*e*_ could be computed as

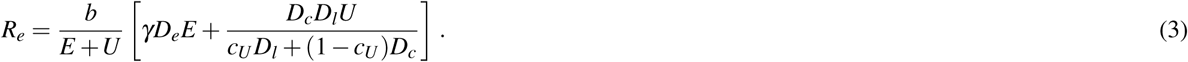

*R*_*e*_ depends on time due to the time dependence of *E* and *U*.

The COVID-19 transmission dynamics (the ODE system) was simulated using the Forward Euler method, with each day discretized into 24 smaller time periods to ensure the numerical stability (see Fig. S3). The parameter fitting was conducted under the Bayesian formulation that combines the effect of the underlying dynamics governed by the ODE system, serving as the prior knowledge, and the collected data, appearing in the likelihood function, to generate the posterior distribution that characterized the behavior of the state variables, including *S, E, I,U, R*, as well as the two unknown parameters, *b* and *r*. For this classical data assimilation problem, we employed the Ensemble Kalman Filter method that was derived from the Kalman filter and tailored to deal with problems with high-dimensional state variables^44,45^. The method proves to be effective when the measuring operator is linear and the underlying dynamics is Gaussian-like. It has been applied to a vast of problems that do not strictly satisfy the Gaussianity requirement. To apply this method, we generated 2000 samples according to the prior distribution, and evolve the samples through the dynamics of the ODE system. The samples were then rectified at the end of each day, using the announced number of confirmed cases, for tuning the two unknown parameters *b* and *r*.

At the beginning of the simulation, March 1, only a few states had non-zero confirmed cases. The true numbers of exposed people and unreported cases on that day, however, are unknown. These two numbers are also the state variables that need to be inferred to using the collected infection data. On March 1, we put a non-informative prior with range [0, 500] and [0, 200] over the exposed latent population and unreported infectious population in each state, respectively. Fig. S4-S13 show the data assimilation results for different states including the number of people in different compartmental groups and their temporal changes with 95% credible intervals. The average reporting rate *r* over all states is 0.2266 at the end of March 20 through the data assimilation method.

For forecasting (in supplementary material), we performed scenario studies of two types. First, we ran the mathematical model by applying the initial data obtained as of March 20 into the future for the next 40 days, but with different configurations of (*b, r, α*_*t*_). The simulation results out of this setting were then compared with those from the setting that the three parameters remained unchanged for each state. To quantify and visualize the difference, we compared the increase of the percentage of the non-affected population when the measures of stay-at-home, increasing test rate, and travel bans were enacted.

The second scenario was about a more ideal situation: every confirmed case would get isolated immediately, as well as those who had been exposed to those confirmed cases, no matter if those who had been exposed had started to show symptoms or not. We built a new mathematical model that incorporated such isolations to study the effect of them. A new quarantined compartment (*Q*) was introduced into the model. Through the simulation, we examined the correlation between the average action-taking time (i.e., temporal lag in putting a person into quarantine denoted by *D*_*q*_) and the increase of non-infected population. In both scenario studies, the simulation was run with the Forward Euler ODE solver, during which each day was divided into 24 intervals to achieve a numerical stability.

As a SEIR-type epidemic model, this model describes the dynamics of different compartments of the population, and assumes homogeneity within each compartment. However, we should note that this assumption may not be valid in real-world scenarios with heterogeneous populations and infections. Indeed, when an individual contracts the disease, the status could be either mild or severe. In our model, this is absorbed by the report rate *r*_*i*_ but is not explicitly differentiated in the model. A more sophisticated model should have the heterogeneities included, but that would pose a significant higher computational demand and more detailed empirical or clinical data support. We leave that to future research efforts.

## Data Availability

The epidemiological data were retrieved from an open source project: Novel Coronavirus (COVID-19) Cases, developed by the Center For Systems Science and Engineering at the Johns Hopkins University (\url{https://github.com/CSSEGISandData/COVID-19/tree/master/csse_covid_19_data}). In addition, we collected millions of points of interest (POIs) with their foot-traffic and anonymous mobile phone users' travel patterns in the United States from SafeGraph. The data for academic research can be requested at \url{https://www.safegraph.com}. The code used for modeling and analysis in this paper is available in the GitHub repository: \url{https://github.com/GeoDS/Travel-Network-SEIR}.

https://github.com/GeoDS/Travel-Network-SEIR

https://github.com/CSSEGISandData/COVID-19/blob/master/csse_covid_19_data/csse_covid_19_time_series/time_series_covid19_confirmed_global.csv

https://github.com/GeoDS/COVID19USFlows

## Data and code availability

The epidemiological data were retrieved from an open source project: Novel Coronavirus (COVID-19) Cases, developed by the Center For Systems Science and Engineering at the Johns Hopkins University (https://github.com/CSSEGISandData/COVID-19/tree/master/csse_covid_19_data). In addition, we collected millions of points of interest (POIs) with their foot-traffic and anonymous mobile phone users’ travel patterns in the United States from SafeGraph. The data for academic research can be requested at https://www.safegraph.com. The code used for modeling and analysis in this paper is available in the GitHub repository: https://github.com/GeoDS/Travel-Network-SEIR/.

## Acknowledgements

We would like to thank the SafeGraph Inc. for providing the anonymous and aggregated human mobility and place visit data. We would also like to thank all individuals and organizations for collecting and updating the COVID-19 epidemiological data and reports.

## Author contributions statement

Research design and conceptualization: Q.L., S.C., S.G.; Data collection and processing: S.C., S.G., Y.H.K.; Mathematical model implementation: Q.L., S.C.; Result analysis: Q.L., S.G., X.S.; Visualization: S.C., S.G., Y.H.K.; Project administration: Q.L. S.G., X.S.; Writing: all authors.

## Supporting Material for

### 1 Mathematical model and configuration

The mathematical model used in this article is derived from the susceptible-exposed-infectious-removed (SEIR) [7, 6, 2, 8] augmented to incorporate human movement, and separate the infected compartment into reported cases and unreported cases. The model is illustrated in the flowchat below:

In the model, for each state, the population is divided into six compartments: *S*_*i*_(susceptible), *E*_*i*_(latent), *I*_*i*_(reported infections), *U*_*i*_(unreported infections) and *R*_*i*_(removed: either recovered or deceased individuals). The subindex *i* stands for the index for the state. Among the six compartments, *S, E* and *U* are “free” people and can move from state to state, while *I*, and *R* are monitored and isolated.

The model is composed of 51 coupled ordinary differential equation (ODE) systems. For each state *i*, the model writes as:

**Figure S1:**
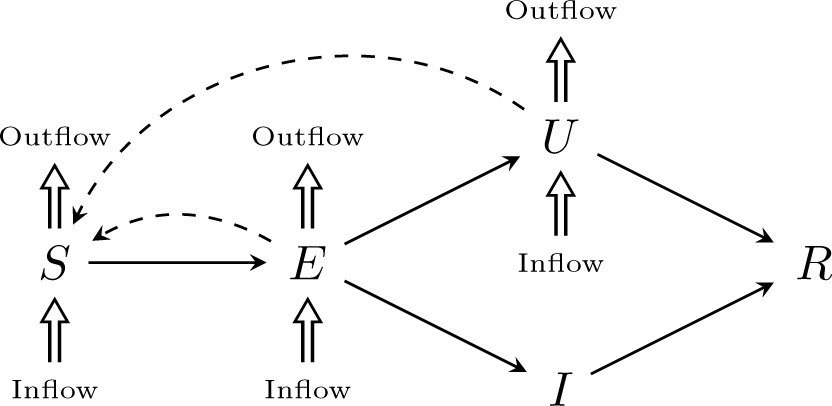
Illustration of the travel flow-network augmented susceptible-exposed-infectious-removed model.

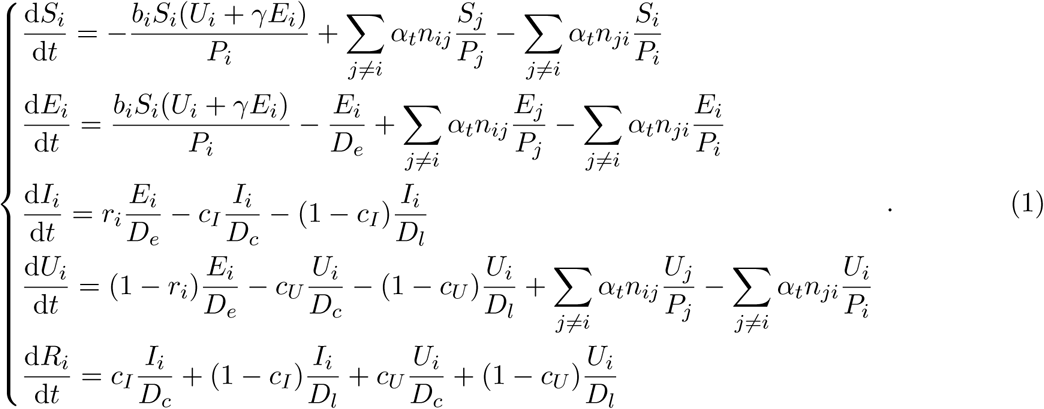

The ODE system is equipped with the following initial data (*t* = 0 standing for March 1, 2020):

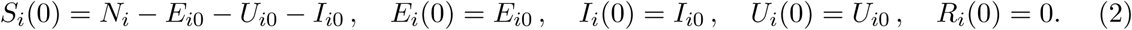

In the equation, the unit for *t* is one day. *N*_*i*_(*t*) is the total population of state *i* at time *t*, and *P*_*i*_ = *S*_*i*_ + *E*_*i*_ + *U*_*i*_ is the free population. *n*_*ij*_ is the number of inflow from state *j* to state *i. b*_*i*_ and *r*_*i*_ are the transmission rate and reporting rate of state *i. c*_*I*_ (*c*_*U*_, resp.) is the proportion of positive cases that show critical condition for *I* (unreported cases *U*, resp.). *D*_*e*_ is the latent period. *D*_*c*_ and *D*_*l*_ are the infectious periods of critical cases and mild cases. *α*_*t*_ is a parameter to tune the traffic flow.

We emphasize two main differences in modeling compared with literature. In [10], the authors study the inter-city traffic and its impact on the spreading of COVID-19 in China. The situation in China and that in the US are very different. In China, the epicenter is clear: the city of Wuhan, Hubei province, and the outbreak starts mid-January, 2020. The COVID-19 outbreak in the US, however, is multi-sourced. The consequence is that in the model in [10], the initial condition for cities excepts Wuhan is clear: the latent, the reported and the unreported cases are all zero. In this model, however, the initial conditions *E*_*i*0_ are unclear for all states; Another big difference is, according to clinical findings, the latent cases also have the potential of transmitting the virus, and thus we add the interaction of *E*_*i*_ with *S*_*i*_ into the increment of *E*_*i*_ [9, 10, 1].

The unknown parameters and state variables in the equation set are

* *b*_*i*_: the transmission rate with non-informative prior range [1, 1.5];
* *r*_*i*_: the report rate with non-informative prior range [0.1, 0.3];
* *E*_*i*0_: the data for the latent population with non-informative prior range [0, 500].
* *U*_*i*0_: the initial data for the unreported population with non-informative prior range [0, 200].
* *S*_*i*0_: the initial data for the susceptible population defined by *N*_*i*_ − *E*_*i*0_ − *I*_*i*0_ − *U*_*i*0_.

Other parameters are:

*γ*: the transmission ratio between unreported and latent. In the simulation we set it to be 0.5;

*D*_*c*_: the average duration of infection for critical cases. We assume *D*_*c*_ = 2.3 days [5].

*D*_*e*_: the average latent period. According to [13], *D*_*e*_ = 5.2 days.

*D*_*l*_: the average duration of infection for mild cases. We assume *D*_*l*_ = 6 days.

*α*_*t*_: the ratio of interstate travel volume compared to that of 2019 during the same period. The travel flow information *n*_*ij*_ was extracted from the SafeGraph mobility data, and we set *α*_*t*_ = 0.5 to represent the travel reduction situation observed in the year of 2020.

*c*_*I*_: proportion of critical cases among all reported cases. We assume *c*_*I*_ = 0.1.

*c*_*U*_: proportion of critical cases among all unreported cases. We assume *c*_*U*_ = 0.2.

There is an essential assumption made in the model: the homogeneity in the population. It means that the traffic flow is a good representation of the total population without considering their demographic and socioeconomic characteristics. The susceptible, exposed, and unreported move in and out of states at the same rate. This explains the 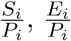 and 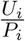 terms in the *S*_*i*_*/E*_*i*_*/U*_*i*_ equation.

The effective reproductive number *R*_*e*_ could be computed as

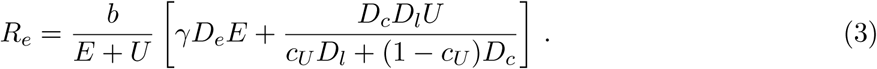

*R*_*e*_ depends on time due to the time dependence of *E* and *U*.

### 2 Data acquisition

#### 2.1 COVID-19 Observation Data

The only available data in the (*S* −*E* −*I* −*U* −*R*) system, for every state, is *I*, the reported confirmed cases. We fetch the data from a community-developed, open source project: Novel Coronavirus (COVID-19) Cases, developed by the Center For Systems Science and Engineering at the Johns Hopkins University [3] ^1^.

#### 2.2 Population and Human Mobility Data

We downloaded the total population data by state in 2019 from the US Census Bureau. In addition, we collected over 3.6 million points of interest (POIs) with travel patterns in the United States from the SafeGraph business venue database^2^. The SafeGraph’s data sampling correlated highly with the United States Census populations^3^. These mobile location data consist of “pings” identifying the coordinates of a smartphone at a moment in time. To enhance privacy, SafeGraph excludes census block group (CBG) information if fewer than five devices visited a place in a month from a given CBG. For each POI, the records of aggregated visitor patterns illustrate the number of unique visitors and the number of total visits to each venue during the specified time window (i.e., March 1st to March 31st 2019 in our dataset), which could reflect the attractiveness of each venue and the national spatial interaction patterns during the last March travel. According to the ODE modeling needs, we further aggregated the travel patterns to the state-to-state spatial scale as shown in Figure S2. In the model, we set the parameter *α*_*t*_ = 0.5 to represent the travel reduction situation observed in the year of 2020 [14].

**Figure S2:**
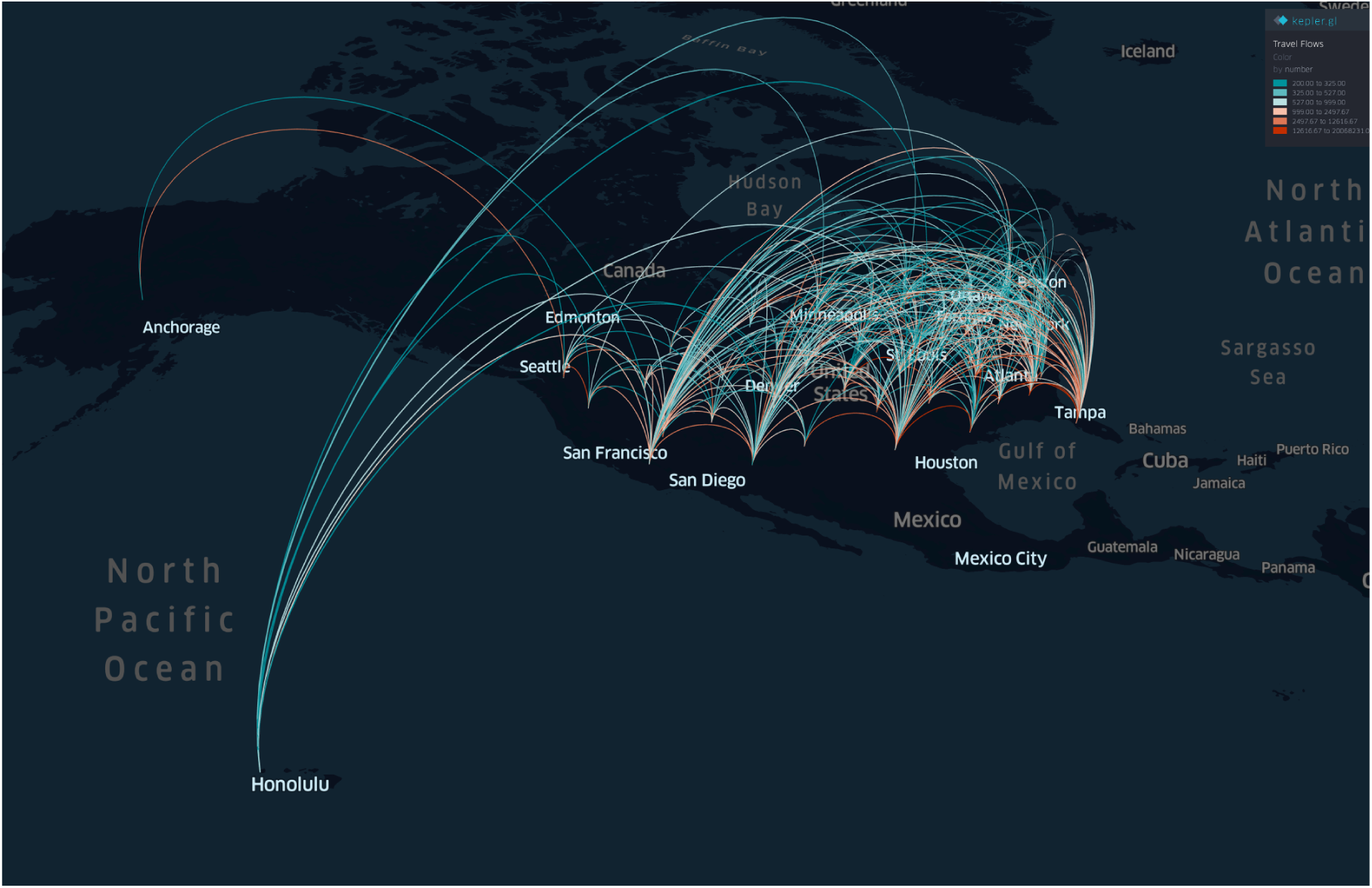
The map of US state-to-state travel patterns aggregated in March 2019 (Data source: SafeGraph; Visualization: kepler.gl)

### 3 Parameter fitting methodology

Each state has its own *S* −*E* −*I* −*U* −*R* data. We assume all the states have their own transmission rate *b*, and the reporting rate *r*. In this section we discuss the method we apply to recover these parameters.

To identify the parameters is a typical data assimilation problem: one has the knowledge from an underlying ODE model, and the access to evolution data. The goal is to build a probability density function that reveals the possible value and the probability of the state variable. Two main ingredients in DA are ODE simulation, and the Bayesian analysis. The ODE system serves as the prior information, and the Bayesian formula blends such dynamical system with the newly fetched data to generate a posterior distribution of the state variables and the parameters. The general flow chart of data assimilation is found in Figure S3.

In the flowchart, *P*_*n*|*n*−1_ is the probability density of *u* upon the evolution step, and *P*_*n*|*n*_ is the probability density obtained through the analysis step.

In our case, *u* is the augmented state variable that includes both the unknown parameters (*b, r*) and the state variable (*I, E, S, U, R*) for every state:

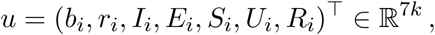

where *k* is the number of states/regions used in the fitting. We transpose it to make it into a column vector. We are aiming at building a distribution density function of *u* over the ℝ^7*k*^ space. 𝒢_*m,n*_(*u*) is the solution to the ODE (1) at time *t*_*n*_ with *u* serving as the parameter in the equation at time *t*_*m*_. The measuring operator is 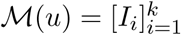, and thus we denote

**Figure S3:**
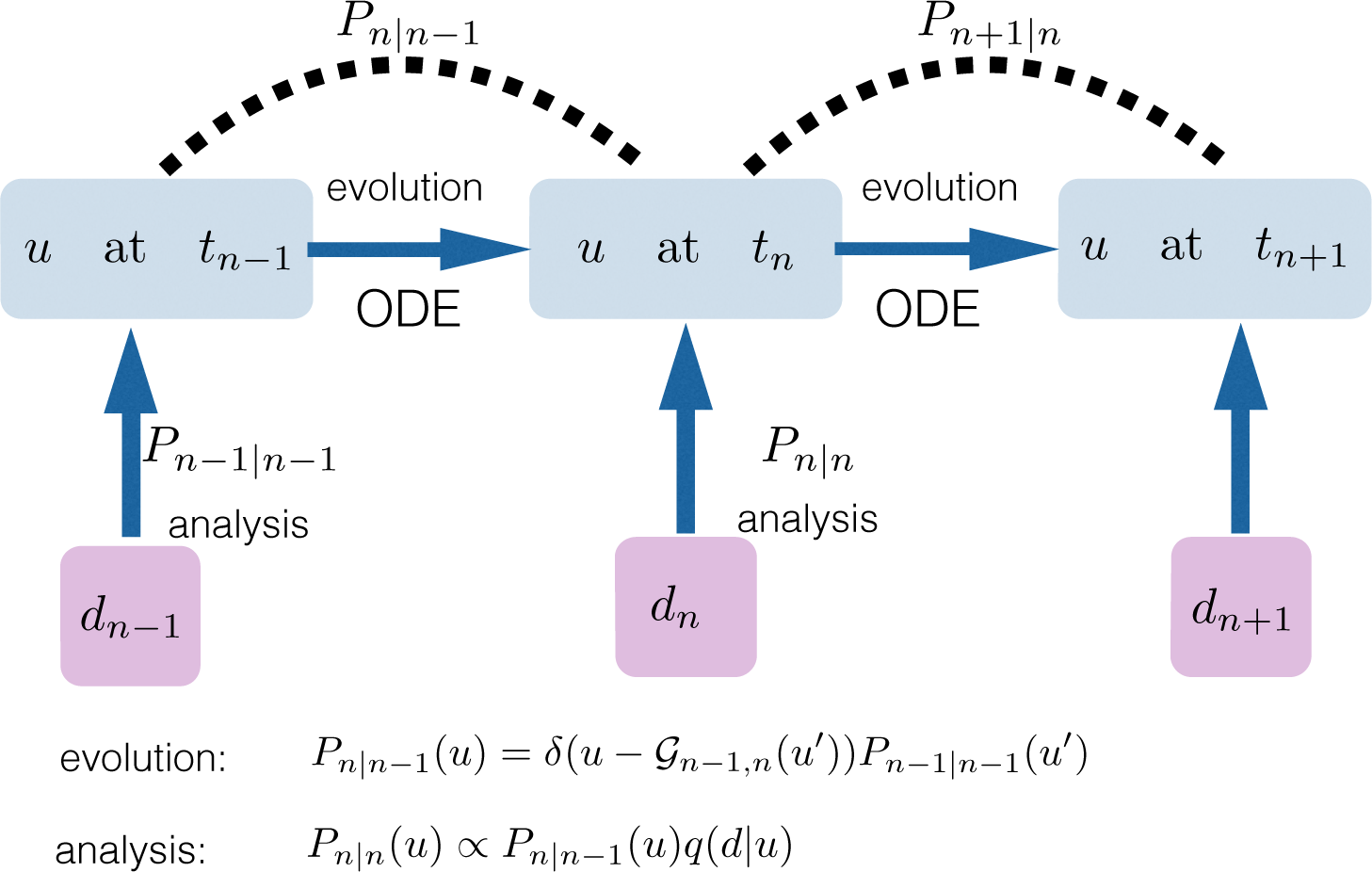
Flowchart of data assimilation: *u* is the state variable, 𝒢_*n*−1,*n*_(*u*) is the forward map by running the ODE from time step *t*_*n*−1_ to *t*_*n*_. *q* is the likelihood function that measures the probability of the error term *d* − ℳ*u* where ℳ is the measuring operator.

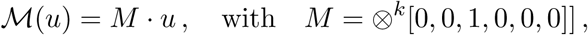

meaning *M* is a 7*k* length vector that has *k* nontrivial entries that pick up all *I*_*i*_ information.

We further assume the collected data has a Gaussian perturbation from the true measuring operator:

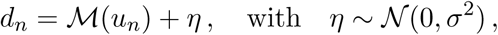

and thus naturally the likelihood function is:

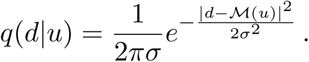

Then together with the two formula for the evolution step and the analysis step:

- Evolution *P*_*n*|*n*−1_(*u*) = *δ*(*u* − 𝒢_*n*−1,*n*_(*u*′))*P*_*n*−1|*n*−1_(*u*′)
- Analysis: *P*_*n*|*n*_(*u*) ∝ *P*_*n*|*n*−1_(*u*)*q*(*d*|*u*)

one can iteratively update *P*_*n*|*n*_(*u*), giving *P*_*n*−1|*n*−1_, the probability density of *u* at time *t*_*n*−1_. In the equation ∝ is the proportional sign: one needs to normalize *P*_*n*|*n*_ to make it a probability density function so that ∫*P*_*n*|*n*_(*u*)d*u* = 1.

There are many choices of data assimilation methods. We choose to utilize Ensemble Kalman Filter that is steered towards analyzing systems having high dimensional state variables. It is a technique, derived from the classical Kalman Filter for the application in atmospheric science, with the analytical covariance matrix replaced by the ensemble version, eliminating the computation of the the Kalman gain matrix and the Riccati Equation that are typically expensive in high dimensional space. It is proved to the effective in the Gaussian case for linear forward model and the measuring operator [4, 11, 12]. The idea is to sample a fixed number of particles on the state variable space according to the initial distribution, and move these particles around at every discrete time step with certain dynamics, to represent the newly adjusted distribution. Denote the number of particles by *N*, and the *j*-th particle after the evolution step at time *t*_*n*_ by 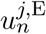, and the *j*-th particle after the analysis step at *t*_*n*_ by 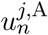, we now summarize the algorithm:

- Evolution:

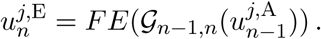

where *FE* is the Forward-Euler discretization applied on ODE (1).
- Analysis:

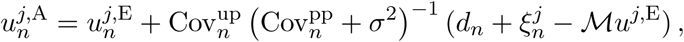

where 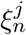 is a *k*-length vector with each entry being random variable i.i.d. drawn from 𝒩 (0, *σ*^2^), *d*_*n*_ is the *k*-length vector collecting the reported infected data on *k* counties. To compute the covariance matrices, we set:

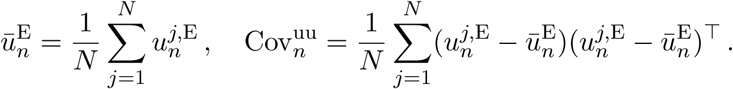

Then naturally

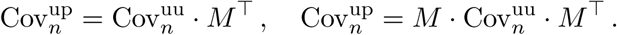

are matrices of size 7*k* × *k* and *k* × *k* respectively.

### 4 Results

The results are divided into two categories: 1. parameter fitting; 2. COVID-19 infection prediction. The computation is done on the state level. Data from 50 states and D.C. (thus *k* = 51) in the United States are used. The model and data assimilation analyses were ran from March 1 to March 20, 2020, and we predict the future infectious cases in different states from March 21 to April 29, 2020 with different parameter setting scenarios.

#### 4.1 State results

For parameter fitting, we utilize the method discussed in Section 3. Total 2000 samples with non-informative prior are adopted to determine 7 × 51 = 357 state variables. The standard deviation of noise is set to be *σ* = 10. In Figure S4-S5, we plot the susceptible, exposed, unreported infections and removed, in time, for the top 10 states with most total confirmed cases as of March 20, respectively. In Figure S6-S7, we plot the reported infections in time, for these 10 states. For most states, the number of reported case grows essentially exponentially fast. Figure S8-S11 show the inferred transmission rate *b* and reported rate *r*. Figure S12-S13 show the time series of effective reproductive number *R*_*e*_ for different states. The signal in the data is rather weak, and for some states, the number of *E* and *U* cannot be inferred at the early stage of the breakout, leading to *R*_*e*_ = 0 for a small period of time for some states.

**Figure S4:**
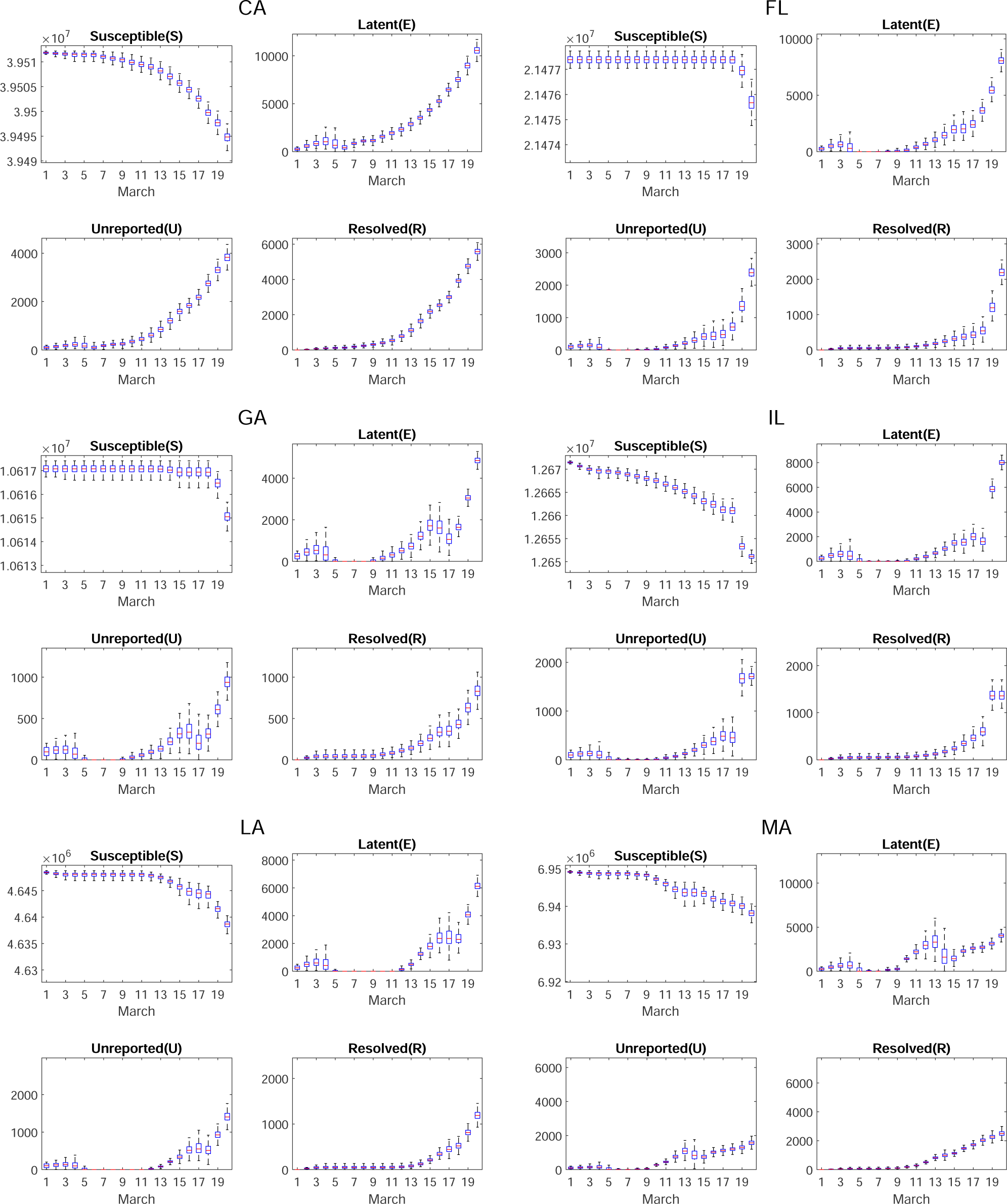
Simulation of *S* − *E* − *U* − *R* for different states. The box and whiskers show the median, interquartile range, and 95% credible intervals.

**Figure S5:**
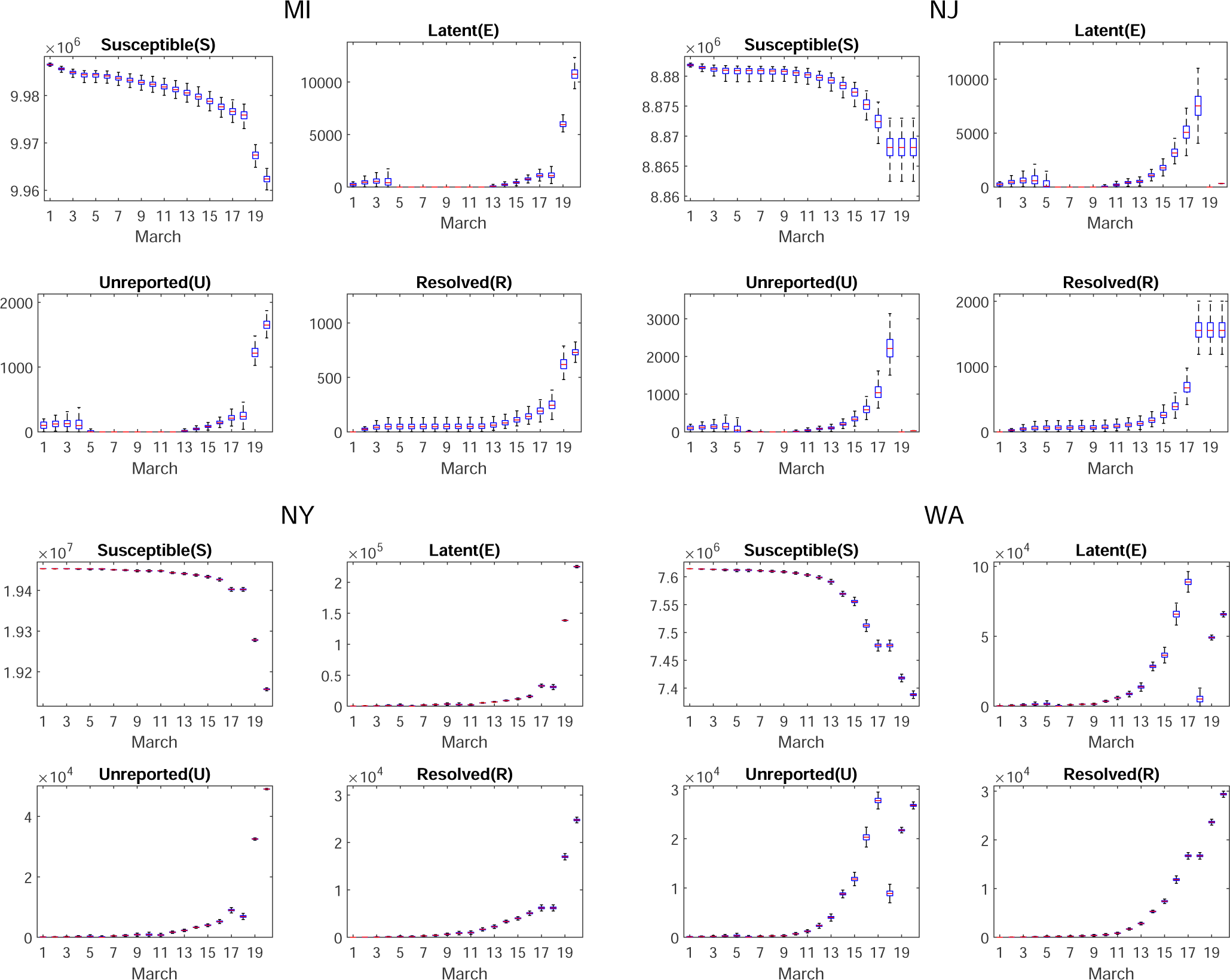
Simulation of *S* − *E* − *U* − *R* for different states. (continued)

**Figure S6:**
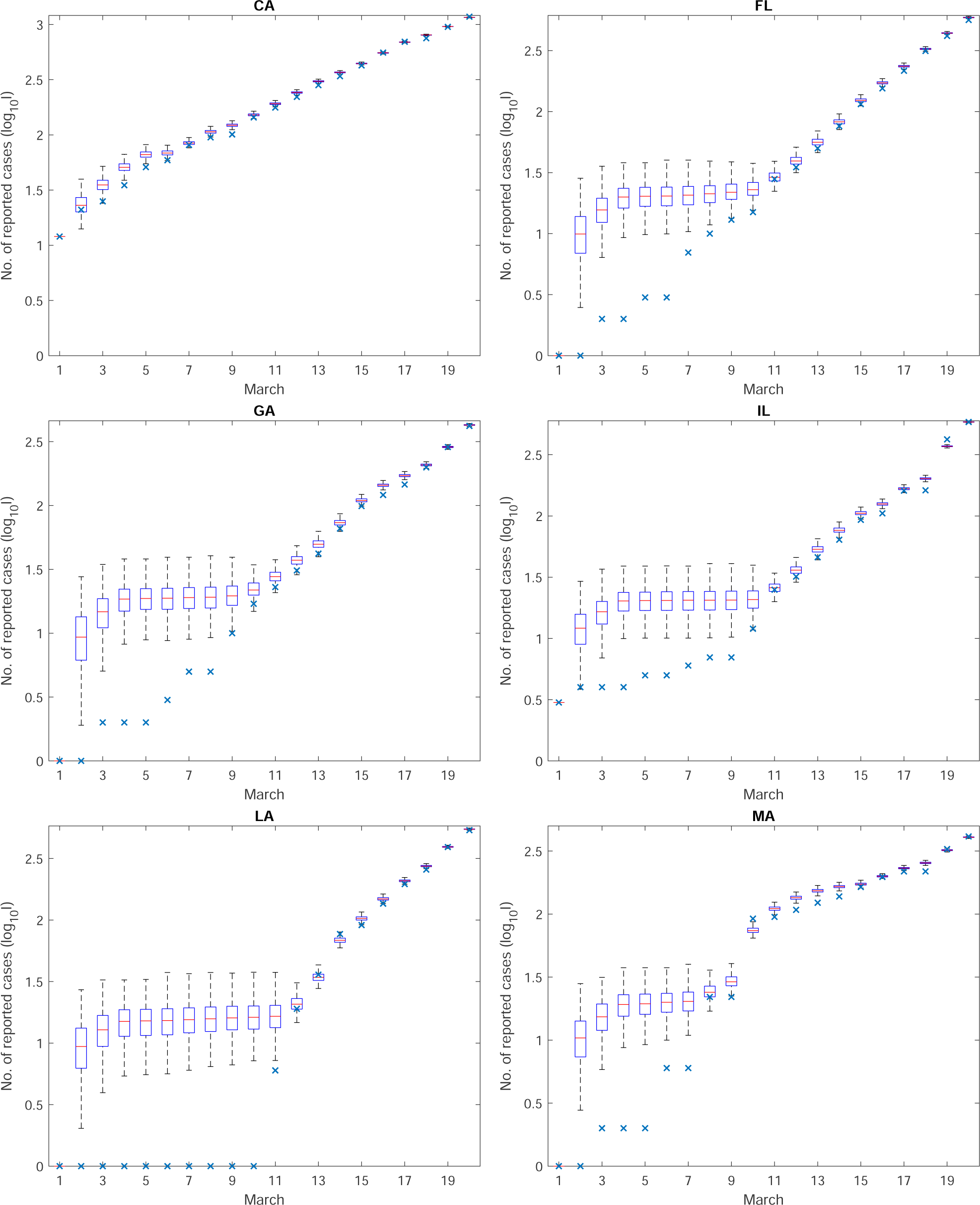
Simulation of confirmed cases *I* (boxes) and the true confirmed cases *I*_true_ (blue x’s) for different states. The box and whiskers show the median, interquartile range, and 95% credible intervals.

**Figure S7:**
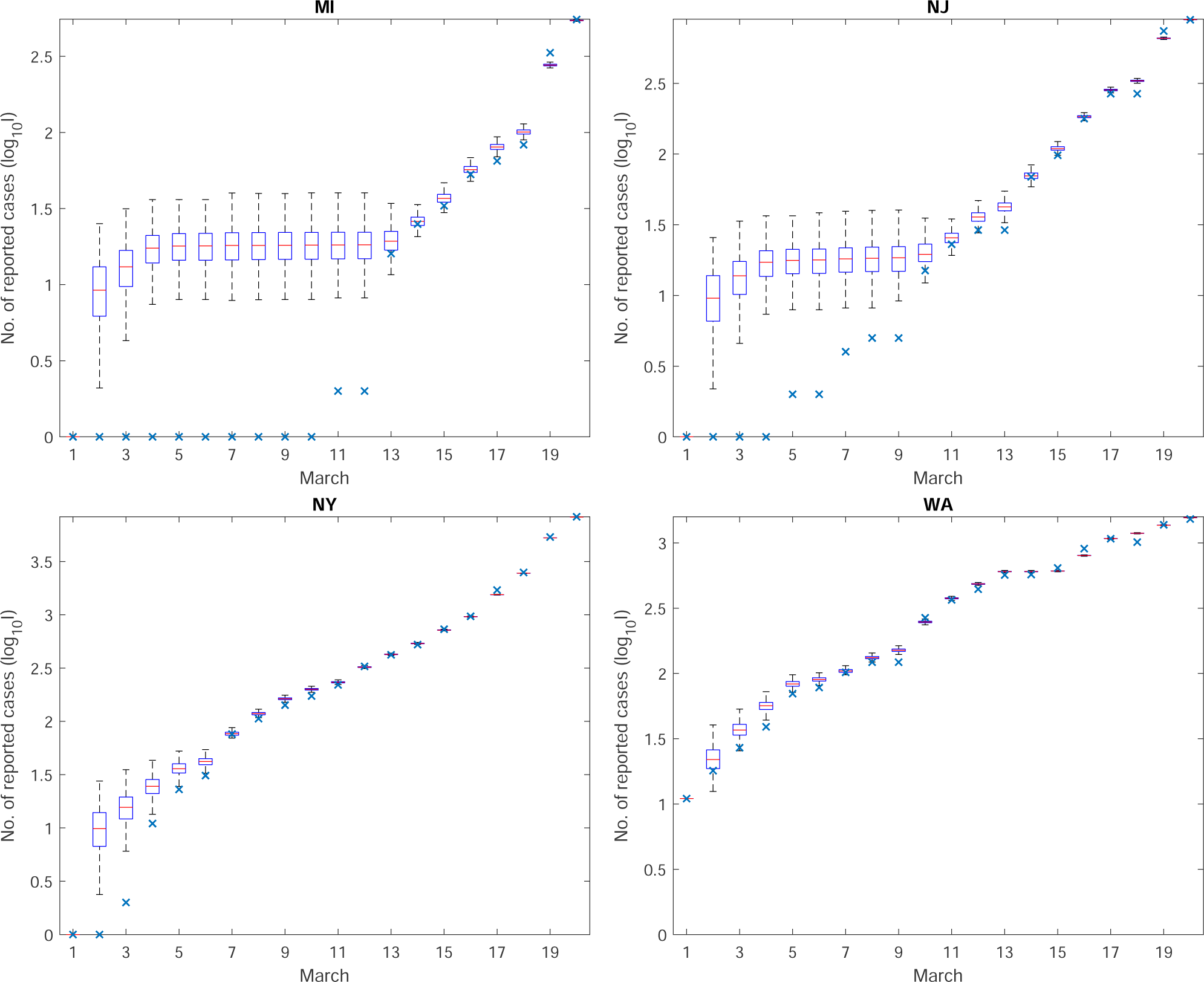
Simulation of confirmed cases *I* (boxes) and the true confirmed cases *I*_true_ (blue x’s) for different states. (continued)

**Figure S8:**
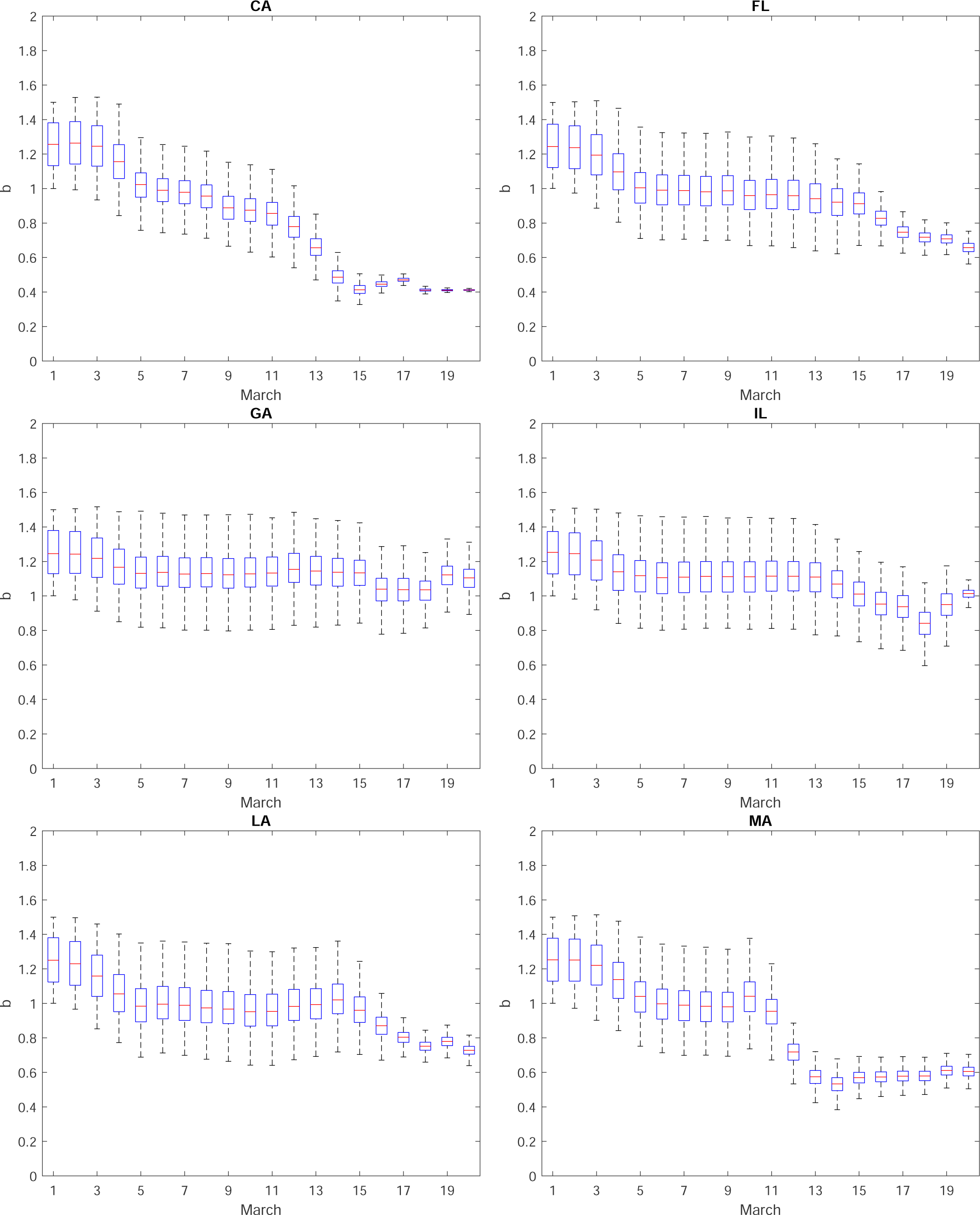
Inferred transmission rate *b* for different states. The box and whiskers show the median, interquartile range, and 95% credible intervals.

**Figure S9:**
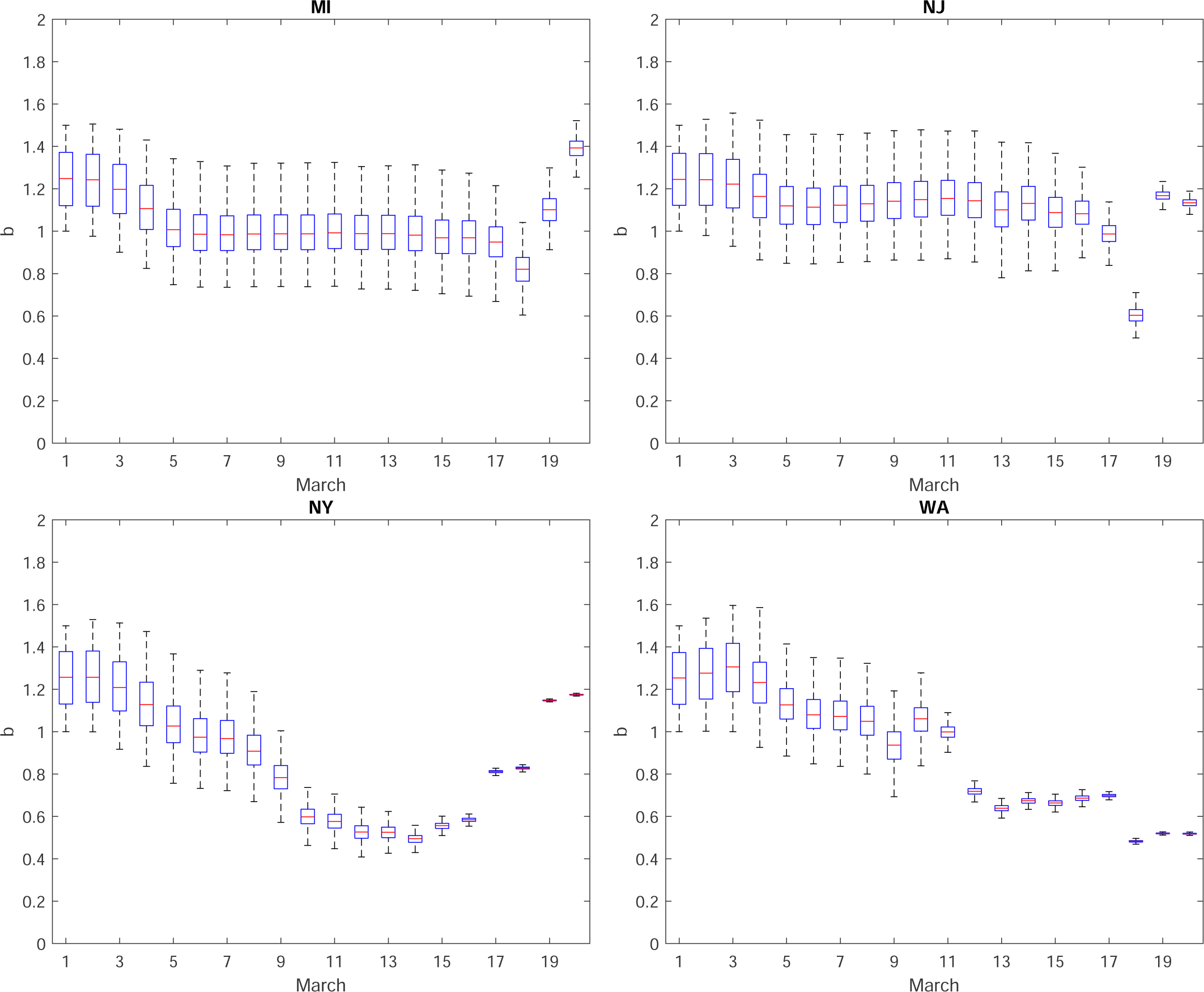
Inferred transmission rate *b* for different states. (continued)

**Figure S10:**
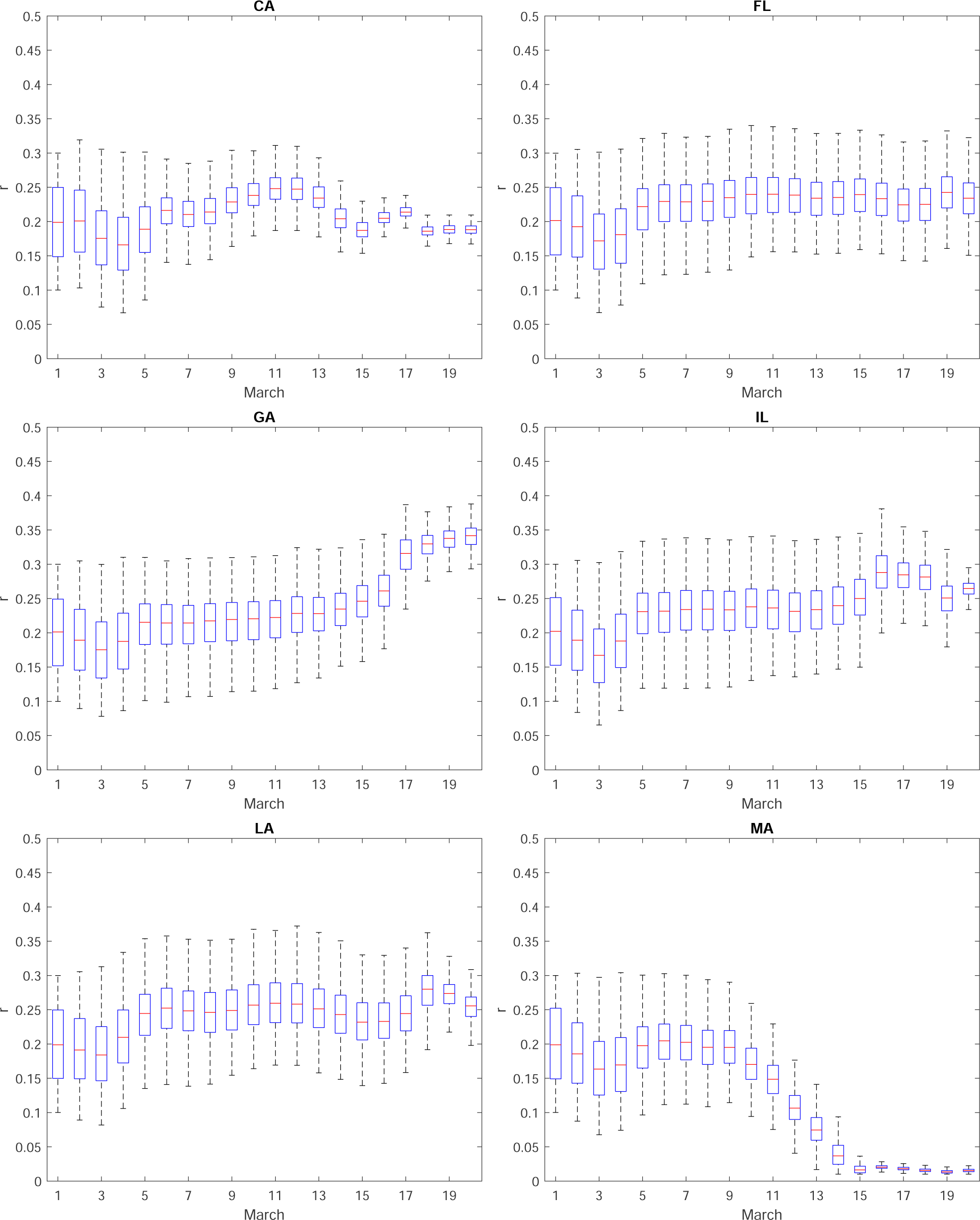
Inferred reporting rate *r* for different states. The box and whiskers show the median, interquartile range, and 95% credible intervals.

**Figure S11:**
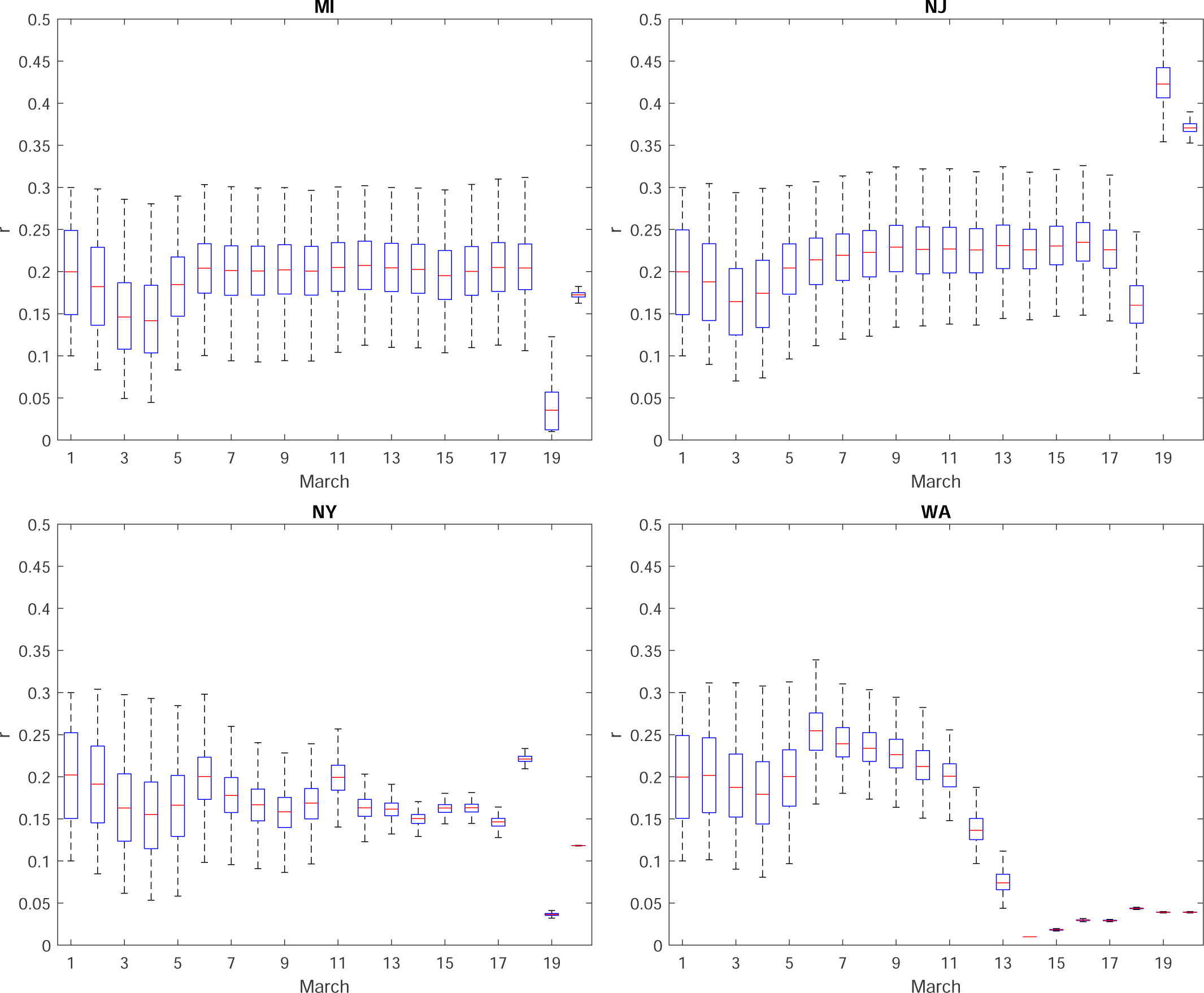
Inferred reporting rate *r* for different states. (continued)

**Figure S12:**
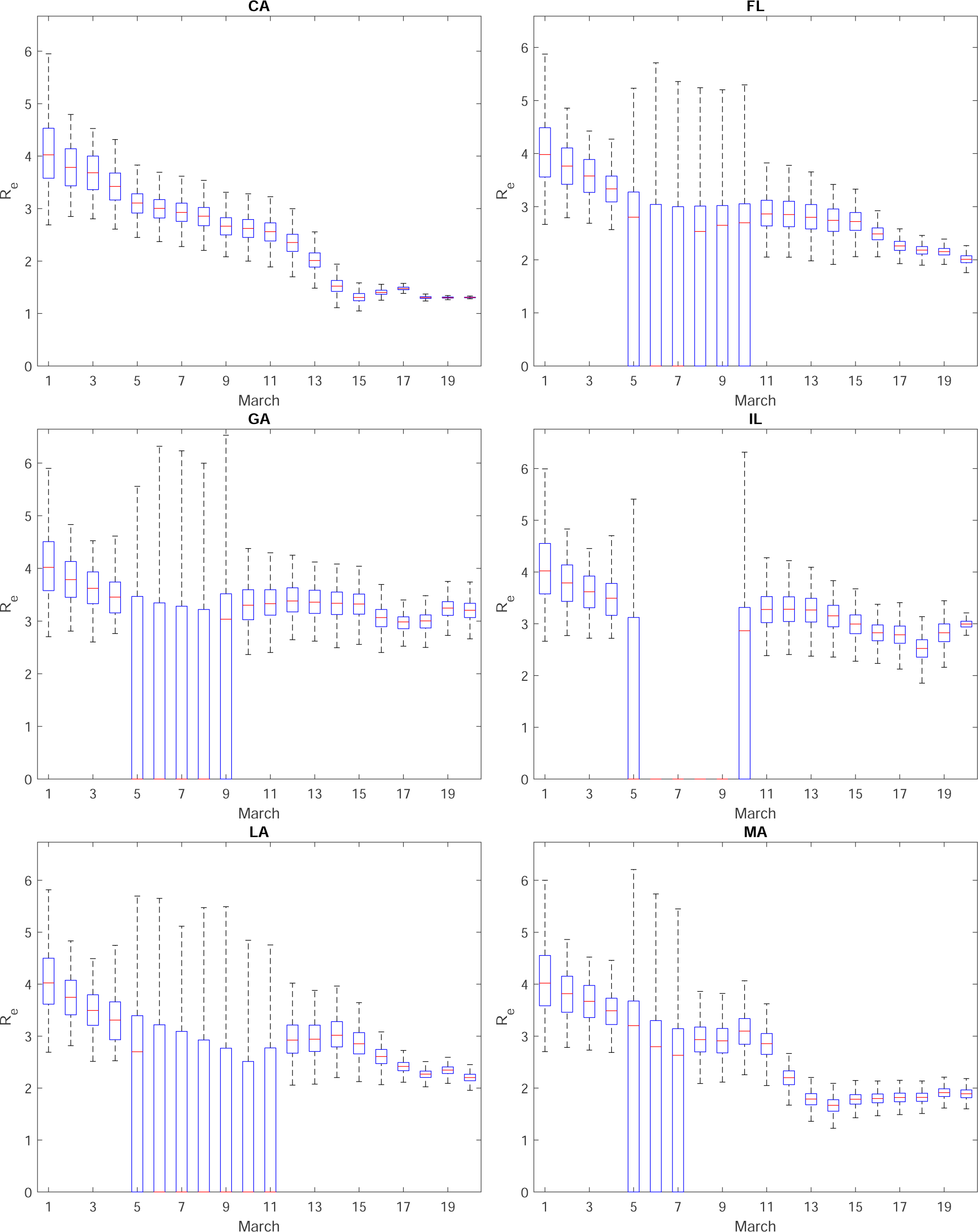
Inferred effective reproductive number *R*_*e*_ for different states. The box and whiskers show the median, interquartile range, and 95% credible intervals.

**Figure S13:**
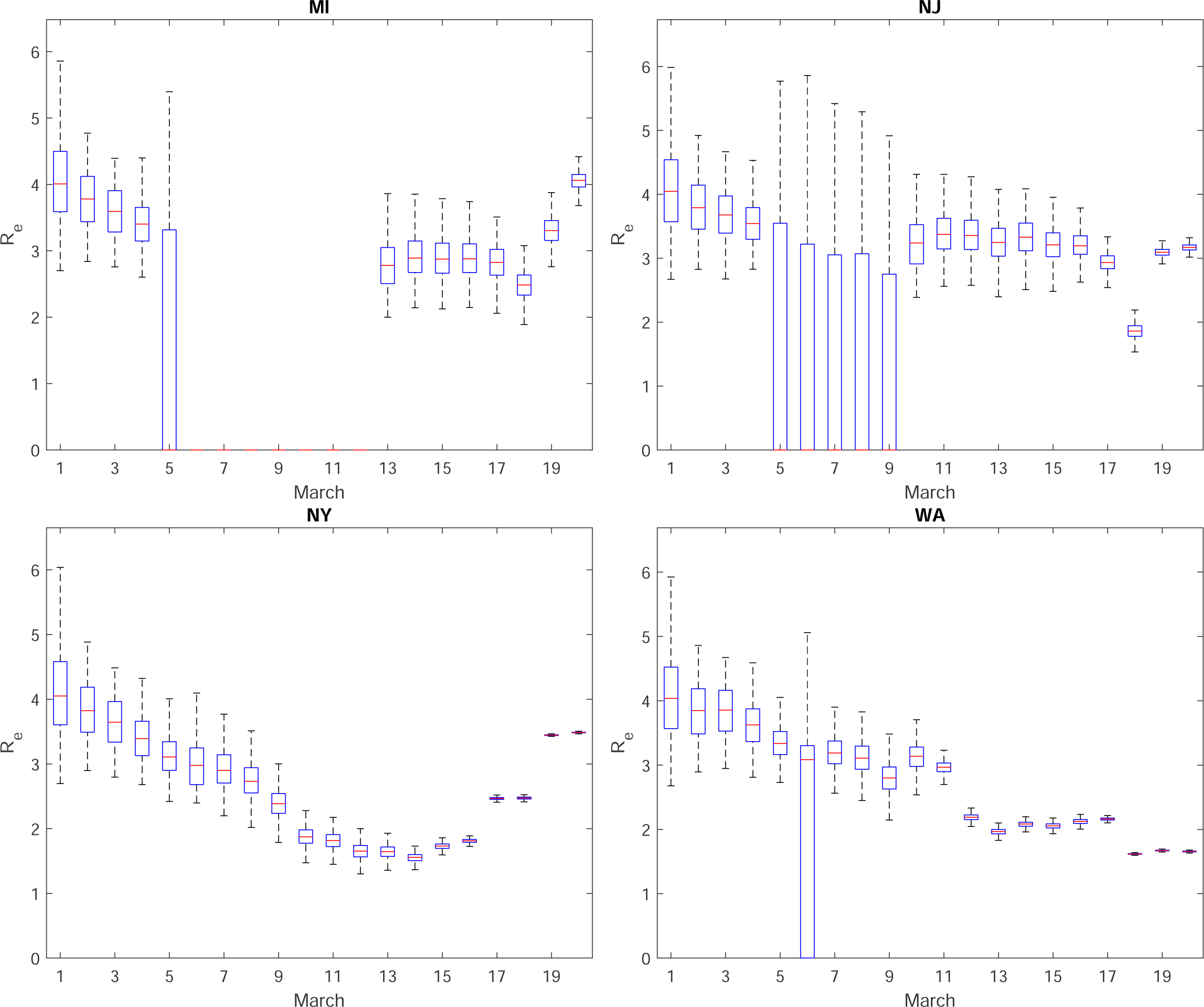
Inferred effective reproductive number *R*_*e*_ for different states. (continued)

#### 4.2 Prediction

The first part of our prediction includes the case study for different transmission rate *b* and reporting rate *r*. In the following, we define the ratio between transmission rate in prediction step and data assimilation step (March 20) by *α*_*b*_, and define the ratio between unreporting rate 1−*r* in prediction step and data assimilation step (March 20) by *α*_*r*_, namely

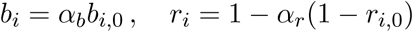

Fig S15 shows the effective reproductive number *R*_*e*_ on April 29 as a function of *α*_*r*_ and *α*_*b*_ for five states.

We further take a proactive approach by directly identify and quarantine the exposed population. The time in days used to discover and isolate them is, on average *D*_*q*_. The model is changed to:

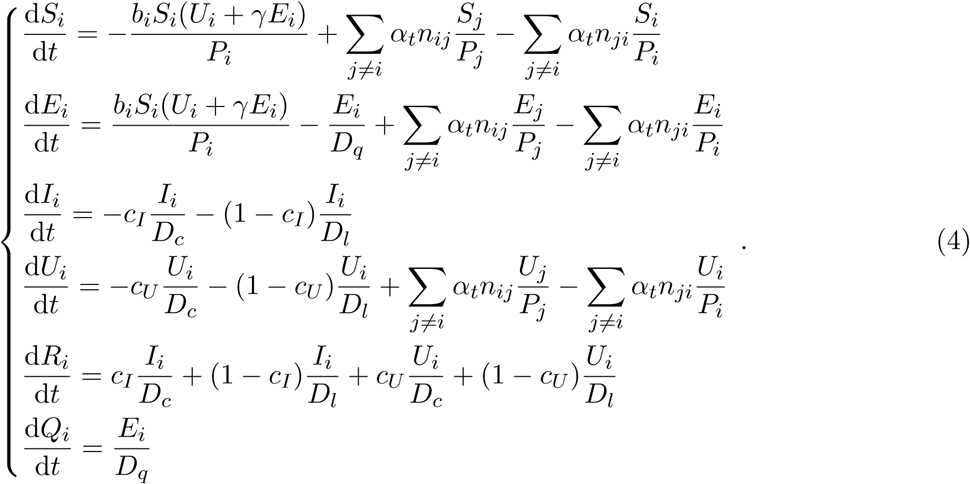

As shown in Fig. 3c, the timely self-quarantine and strict isolation right away or starting at most about 3.6 days (the median value) after exposed to the SARS-CoV-2 virus is found most effective in containing the COVID-19 outbreak for most contagious states in the US.

#### 4.3 Limitation

For the scope of this paper, we do not consider the hospital capacity, and thus we assume the current or timely built facility can accommodate all reported infected cases. However, if one can have access to the hospital capacity data in each state, the capacity information can be fed to the model as well. Furthermore, we do not distinguish mild and severe symptoms in the model.

## Footnotes

1 https://github.com/CSSEGISandData/COVID-19/blob/master/csse_covid_19_data/csse_covid_19_time_series/time_series_covid19_confirmed_global.csv

2 www.safegraph.com

3 ^3^www.safegraph.com/blog/what-about-bias-in-the-safegraph-dataset

